# Nasopharyngeal aspirates vs. nasal swabs for the detection of respiratory pathogens: results of a rapid review protocol

**DOI:** 10.1101/2020.10.21.20216077

**Authors:** Matthew F. Flynn, Martin Kelly, James S. Dooley

## Abstract

**Background:** Nasal pathogen detection sensitivities are often as low as 70% despite advances in molecular diagnostics. It has been suggested that this is linked, in part, to the choice of sampling method.

**Methods:** A diagnostic test accuracy review for sensitivity, using recently developed Cochrane methods for conducting rapid reviews, and the PRISMA protocol was undertaken, with QUADAS-2 risk of bias assessments and meta-analysis of included studies. Sensitivities were calculated by a consensus standard of positivity by either method as the gold standard. Insufficient and/or inaccurate, cross sectional or anatomical site pooling methodologies were excluded.

**Results:** Of 13 included studies, 8 had ‘high’ risk of bias, and 5 had ‘high’ applicability concerns. There were no statistical differences in pooled sensitivities between collection methods for 8 different viruses, and neither with use of PCR, Immunofluorescence nor culture. In a single study, Influenza H1N1 favoured nasopharyngeal swabs, with aspirates having 93.3% of the sensitivity of swabs (p>0.001). Similar equivocal sensitivities were noticed in detecting bacteria.

**Conclusions:** The chain of sampling, from anatomical site to laboratory results, features different potential foci along which sensitivity may be lost. A sufficient body of evidence exists that use of a different sampling method will not yield more respiratory pathogens. The new Cochrane Rapid Reviews guidance helped rapidly answer this relevant and timely clinical question.

## Background

Accurate laboratory-confirmed diagnoses aid both timely treatment and surveillance of respiratory infections at global levels, through rapid detection^123^. The frustration of false negative results for specific pathogen carriage, experienced by clinicians thus escalating treatment upon clinical suspicion alone, predates SARS-CoV-19, and leads to reliance on repeat tests and imaging^45^. Suboptimal sensitivity has persisted for viruses despite the adoption of Polymerase Chain Reaction (PCR) as the gold standard above viral culture and direct immunofluorescence (DIF), where increased specificity of PCR has in itself decreased sensitivity in comparison to other methods due to false positives no longer being incorporated^6^. PCR avoids the decreased sensitivities in patients over 5 years of age seen with DIF, and is further advantaged in being able to quantify viral load (qPCR)^789^. Higher viral loads present in the early course of a viral infection predominate in the nose than in the throat, and slightly predominate in the nasopharynx than the anterior nasal cavity^10,11,12^. For bacteria, there is a related but distinct microbiome between the anterior nares and nasopharynx^13^. The nasopharynx is the uppermost portion of the throat lying at the back of the nasal cavity and accessible horizontally along the nasal floor past occasionally obstructing turbinates and deviations of the septum. A nasopharyngeal swab (NPS) is inserted to a depth equal to the distance from the nostril to the earlobe (nasotragal length, NTL), or until the nasopharynx is felt (a depth of up to 14cm), with less deep swabs >=5cm sampling the middle meatus or anterior nares^14,15,16,17^. The NTL in children and infants is shorter, but remains considerably longer than the 2cm depth occasionally cited in NPS studies, and is well described in approaches to paediatric intubation^18,19^. Lack of clarity over such definitions and ensuing heterogeneity has frustrated meta-analysis of this area^20^. A review of methods for Influenza detection found increased yield when pairing combinations of diverse methods^21^. Combined oropharyngeal and anterior nasal swabs are shown to be comparable in sensitivity to a single sample of the nasopharynx whilst benefiting from higher patient satisfaction^22,23,24^. These combined throat/nose swabs have become recommended practice for self-administration of the test^25^. Paired oropharyngeal/NPSs convey increased sensitivity compared to NPS alone^26,27^. The swab type used is an important consideration, with greater yield of respiratory epithelial cells and greater patient satisfaction with a flocked swab (akin to a miniature toilet brush) than a rayon-tipped swab (resembling a long cotton “ear” bud) but pathogen detection rate is equivocal^28,29^. Other important pre-laboratory variables such as pre-impregnation of swabs with transport media, immediate placing in medium following collection or refrigeration of the sample appear to add little to the diagnostic yield^30,31^. Nasal aspiration (NA) involves the removal of mucous from the nasal cavity with a suction catheter, which is then removed with a mucous trap and subsequently flushed with saline or transport medium. The similar but distinct nasal wash (NW), also called the Naclerio method, is obtained by the drainage without suction of a small volume of saline flushed into the nose^32,33^. Samples obtained by nose blowing are not widely used, and the high prevalence of *Staphylococcus aureus* in these samples suggests microbial contamination from more keratinised topographies than just the internal nose^34,35^.

Whether or not a paired test is used, whether tests are repeated, and whether or not other variables exist, it remains a potent question as to which is the optimal nasopharyngeal sampling method – aspiration, washing or swabbing. In the absence of an objective, categorical gold standard, a dual composite reference standard combining these two imperfect tests can be used to create a “consensus standard” or “either positive” rule, against which to compare sensitivities^36,37^. Such a reference standard of course overestimates true sensitivity, with even 100% sensitivity representing an optimal fraction achievable by that limited method, but allows a head-to-head comparison of the two techniques with eyes open to this innate underestimation of viral presence^38^. This approach has the failing of having one-sided false negatives i.e. all the pathogens detected in one sample were also detected in the other, which thus has a potentially misleading sensitivity of 100%. This issue can be addressed through risk of bias assessments. If mild increases in pathogen yield suffice to influence best practice, then any similar small gains by changing collection method type require analysis of available evidence^39,40^. The current true sensitivity of nasopharyngeal swabbing is estimated at 71-89%^41^. An increase in sensitivity from 70-90% is enough to decrease by more than half the pre-test probability of a true infection up to which it would be deemed safe to act as if a negative test was indeed so^42^. This rapid review is designed to compare the available evidence per pathogen, laboratory method and collection method using new streamlined systematic review methods^43,44^.

## Methods

This systematic review was conducted along the Interim Guidance from the Cochrane Rapid Reviews Methods Group for conducting rapid reviews, following the Preferred Reporting Items for Systematic Reviews and Meta-Analyses (PRISMA) statement, and with reference to the Cochrane handbook for diagnostic test accuracy reviews^45,46,47^. The protocol was registered with the International Prospective Register of Systematic Reviews (PROSPERO) (registration no. CRD42020189577) prior to formal screening of search results against eligibility criteria^48^. The protocol was developed in consultation with knowledge users in respiratory medicine, paediatrics, microbiology and otolaryngology as well as a focus group with members of the public to promote fitness for purpose.

### Inclusion criteria

All studies comparing sensitivity in microbiological sampling for the upper respiratory tract were included in the search. Variations in viruses, bacteria and fungi detected and differing laboratory techniques were included and stratified by these categories in the reported results. The populations included healthy patients, those with respiratory disease (including inoculation of healthy patients), infants populations to adults, and studies in different seasons and climates. These however were not compared, being considered to influence carriage and detection rates, but not influence sensitivity discriminately between collection methods. Studies were limited to those in humans, and those since publication of WHO guidelines for the collection of human specimens for laboratory diagnosis of avian influenza infection (2005)^32^. Pooled samples such as swabbing of the nose and throat by the same swab or where samples were not paired from the same patient were excluded. Where a consensus standard was not derivable from the data, and this information could not be obtained, these studies were also excluded.

### Nasopharynx definition

For the purposes of this review, swabs of the nasopharynx were defined as swabbing to depth of >5cm in adults and >3cm in children or with relevant reference to surface anatomy, by following WHO guidelines, or citing other well documented studies in the methods section. Where this data was lacking, the authors were contacted for clarification, and/or judgments made on inclusion based on use of appropriate anatomical nomenclature and documented training of staff. Broad interpretation of the term “nasopharynx” elsewhere in the literature to include the middle meatus and anterior nares, led to the inclusion of ‘nasopharyngeal’ studies with sparse methodological detail, but graded as ‘high’ for applicability concerns.

### Search strategy

Literature search strategies were developed with a life sciences librarian. Cochrane CENTRAL, MEDLINE and Embase were searched, followed by supplemental exploration of reference lists of other reviews for focused optimization of search saturation. In the interests of time, other databases and grey literature were not included, and studies were restricted to those in English. Peer review of search strategy was not undertaken. The search was conducted on 09/06/2020.

### Study selection

A title and abstract screening form was piloted using 30 abstracts and adopted without modification to dual screen 20% of abstracts with conflict resolution. Remaining abstracts were screened by one reviewer (MF) and the second reviewer screened all excluded abstracts. A full text screening form was piloted using 5 full text articles with the same process followed as title and abstract screening. Web based review portal Rayyan QCRI was used to streamline the selection process while allowing screening to be conducted in parallel^49^. Search results are summarized in the PRISMA flow diagram [figure 1] and search terms are accessible via the registered protocol on PROSPERO.

**Figure 1.**
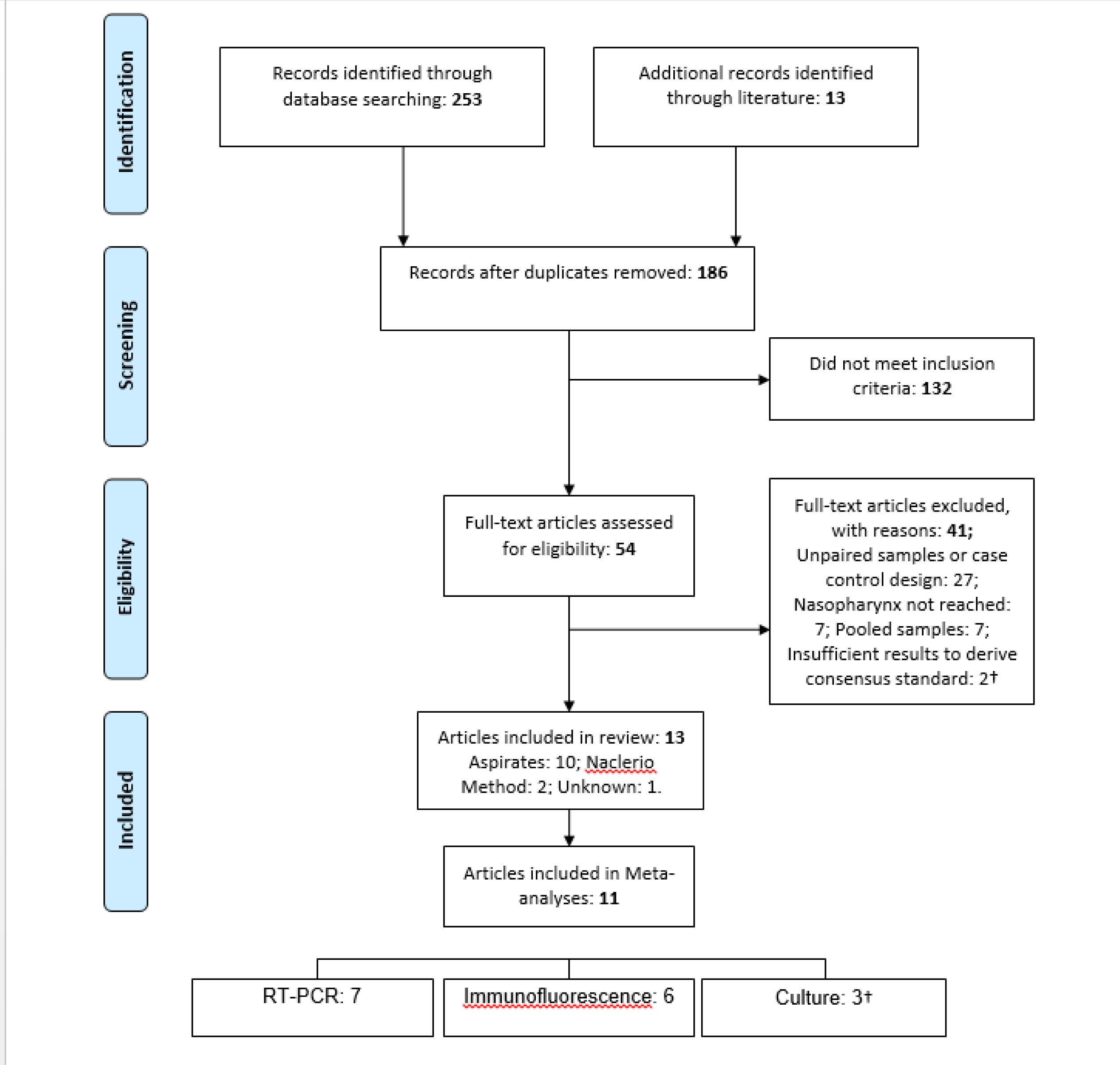
PRISMA Flow chart. RT-PCR: Reverse Transcriptase Polymerase Chain Reaction † Studies may fall in more than one category

### Data Extraction

The following data were collated onto a Microsoft Excel spreadsheet: Publication year, number of participants, collection type, separate nostrils used, age of population, transport medium, analysis technique, swab type and type and quantity of microbiota detected. Variations in viruses, bacteria and mycobacteria and differing laboratory techniques were included and stratified by these categories in the report. Over and above the two designated methods described by the WHO of aspirates (extraction of fluids by suction catheter) and washes (free drainage without suction of instilled saline into a receiving bowl), there was a third hybrid method, where flushed water was then aspirated^32^.” For the purposes of this study, aspirates obtained after instilling water into the nose and those aspirated “dry” have been grouped together under “Aspirate”, and washes not involving suction termed “Washes.” Where studies included other collection methods, compared sensitivities from multiple anatomical sites, or compared sensitivities of different laboratory techniques all calculated with against a gold standard from one collection method alone, only data meeting the inclusion criteria above and demonstrating a head to head analysis of NP swabs and nasal aspirates/washes were extracted. In cases where the consensus standard was equal to the sensitivity of one of the sampling methods i.e. there were no false negatives for this collection method, this data was still included but acknowledged as having ‘high’ risk of bias under the reference standard domain.

### Data Analysis

Where there were a sufficient number of studies, results formed a proportional meta-analysis, else a narrative was used. Sensitivity analysis and McNemars’ test for paired samples were derived using Medcalc and Scistat online statistical software respectively^50,51^.

### Risk of Bias assessment

The QUADAS-2 risk of bias tool for diagnostic test accuracy reviews was used to grade risk of bias and applicability concerns by one reviewer (MF) with verification by second viewer (JD)^52^. This was modified to include the signalling questions “Were false negatives two-sided?” under reference standard, and “Were separate nostrils used?” under flow and timing. Risk of bias tables were collated using RevMan^53^. Studies funded by, or with material help from the company supplying the tests were noted.

## Results

The initial search identified 253 articles [figure 1]. 13 were added from searching reference lists of key papers and reviews. After screening titles and abstracts, the full texts of 186 publications were reviewed. Of the 54 of these which met the eligibility criteria, 13 were included in the final review, including three from supplemental searching and one published abstract. No studies detected fungi or mycobacteria. One study was an abstract published within a conference proceedings supplement.

### Risk of Bias assessment

Using the QUADAS-2 tool, overall risk of bias was high: 8/13 studies with “high” risk of bias. Applicability concerns were “low” in 8/13 studies [figure 2, 3]. Lack of information on the patient selection process in 11/13 led to “unclear” risk of bias under patient selection. There were “low” applicability concerns due to patient selection, but this was a reflection of the review question including any and all populations, there was considerable homogeneity of age, ethnicity or disease status. The high frequency of false negatives being one sided (8/13), led to high risk of bias under the reference standard domain. Three studies declared material support from companies manufacturing the testing kits.

**Figure 2.**
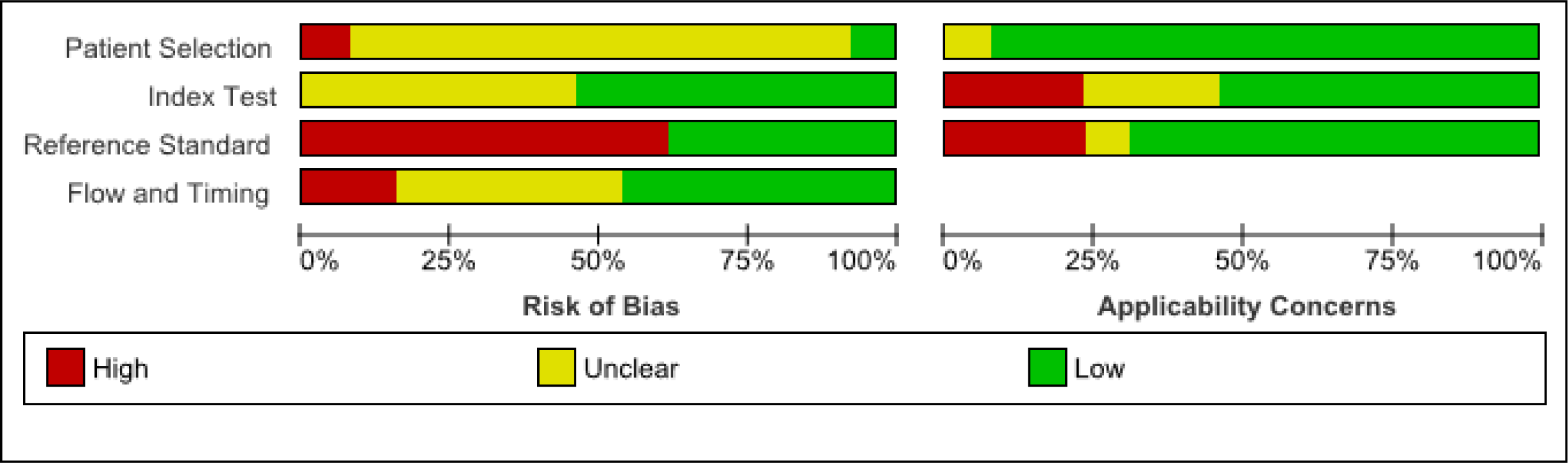
Methodological Quality Summary Chart.

**Figure 3.**
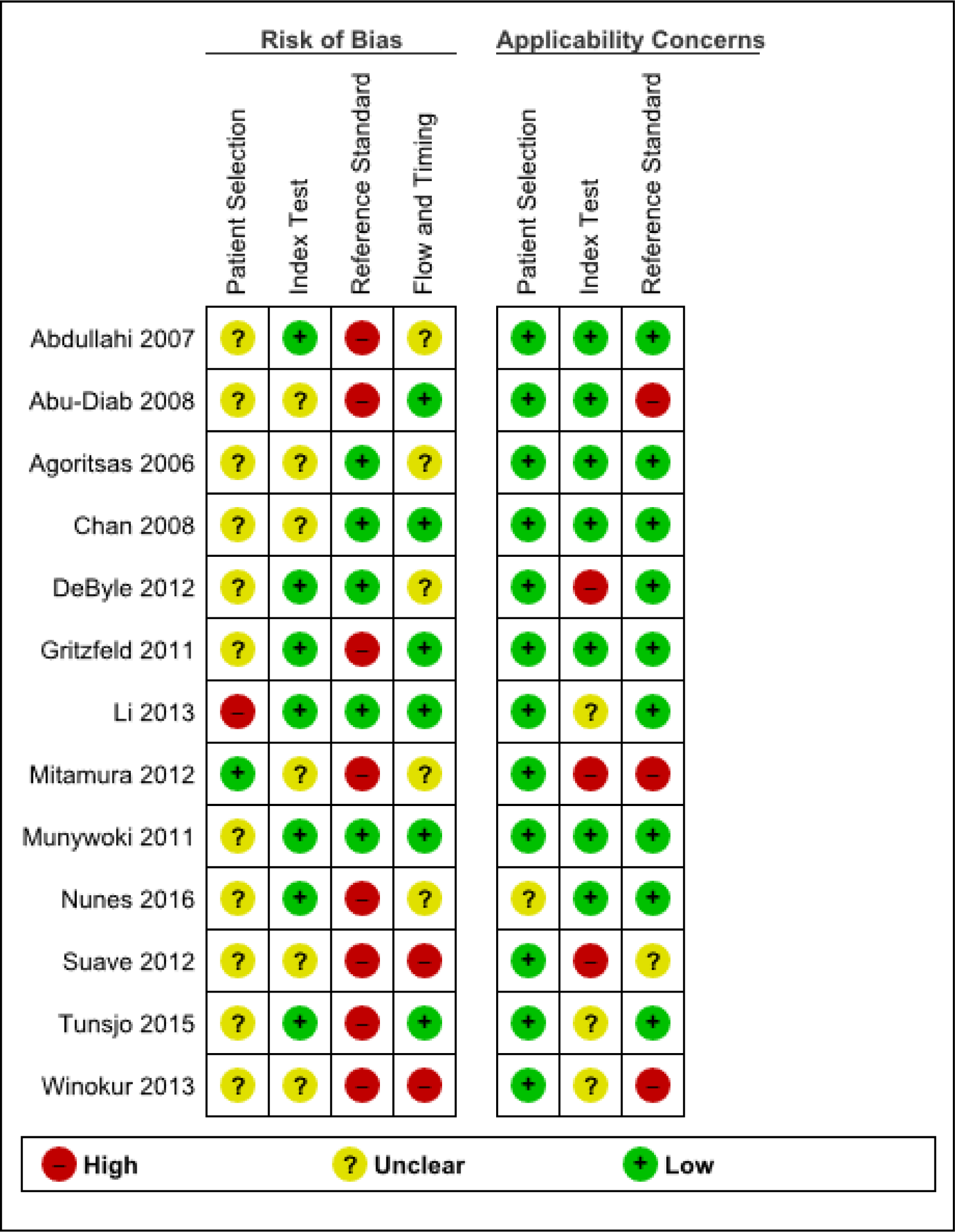
Methodological Quality Summary Table.

### Heterogeneity

The available literature was limited by methodological heterogeneity of participant age, participant health, laboratory methods and collection methods, even within collection methods given the same name [table 1].

**Table 1.**
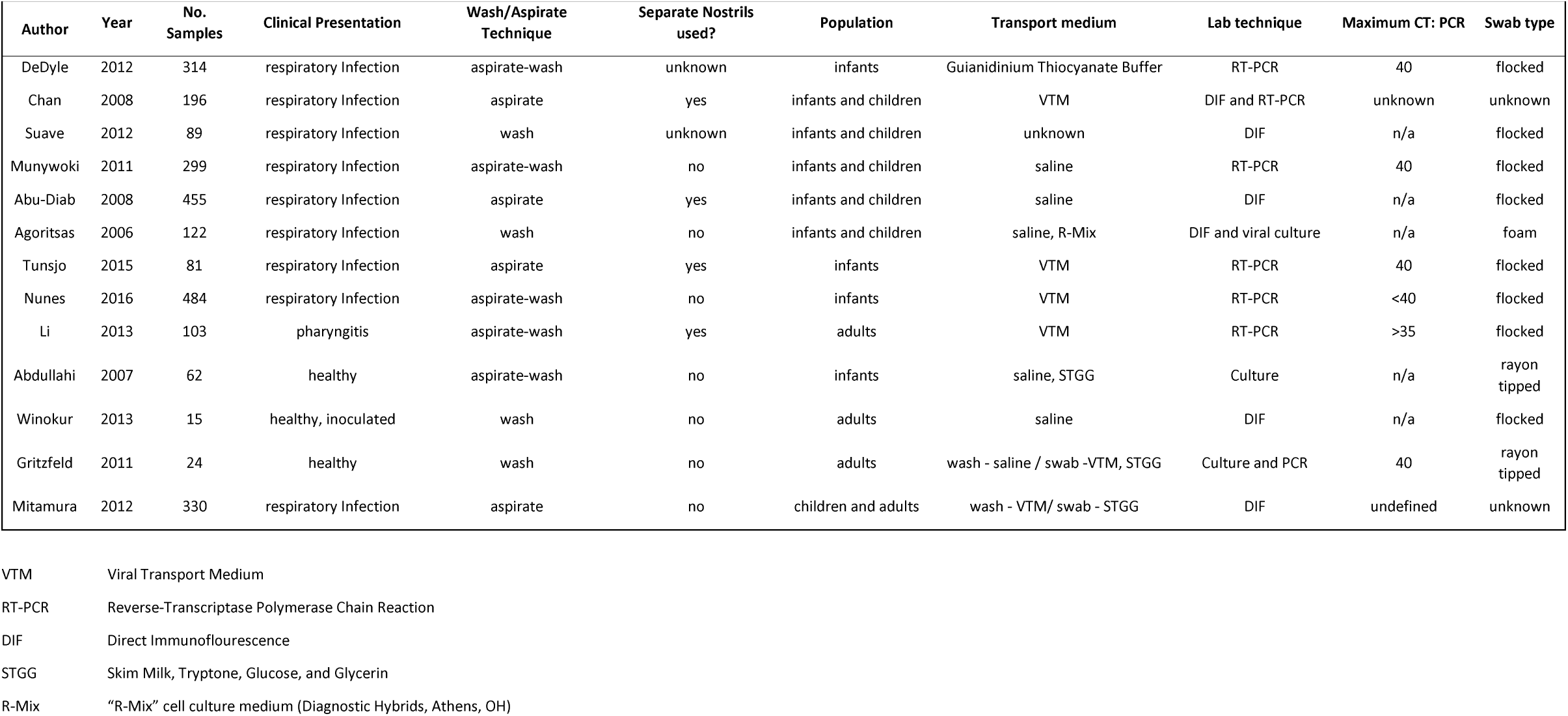
Characteristics of Included studies

| Author | Year | No. Samples | Clinical Presentation | Wash/Aspirate Technique | Separate Nostrils used? | Population | Transport medium | Lab technique | Maximum CT: PCR | Swab type |
| --- | --- | --- | --- | --- | --- | --- | --- | --- | --- | --- |
| DeDyle | 2012 | 314 | respiratory Infection | aspirate-wash | unknown | infants | Guianidinium Thiocyanate Buffer | RT-PCR | 40 | flocked |
| Chan | 2008 | 196 | respiratory Infection | aspirate | yes | infants and children | VTM | DIF and RT-PCR | unknown | unknown |
| Suave | 2012 | 89 | respiratory Infection | wash | unknown | infants and children | unknown | DIF | n/a | flocked |
| Munywoki | 2011 | 299 | respiratory Infection | aspirate-wash | no | infants and children | saline | RT-PCR | 40 | flocked |
| Abu-Diab | 2008 | 455 | respiratory Infection | aspirate | yes | infants and children | saline | DIF | n/a | flocked |
| Agoritsas | 2006 | 122 | respiratory Infection | wash | no | infants and children | saline, R-Mix | DIF and viral culture | n/a | foam |
| Tunsjo | 2015 | 81 | respiratory Infection | aspirate | yes | infants | VTM | RT-PCR | 40 | flocked |
| Nunes | 2016 | 484 | respiratory Infection | aspirate-wash | no | infants | VTM | RT-PCR | <40 | flocked |
| Li | 2013 | 103 | pharyngitis | aspirate-wash | yes | adults | VTM | RT-PCR | >35 | flocked |
| Abdullahi | 2007 | 62 | healthy | aspirate-wash | no | infants | saline, STGG | Culture | n/a | rayon tipped |
| Winokur | 2013 | 15 | healthy, inoculated | wash | no | adults | saline | DIF | n/a | flocked |
| Gritzfeld | 2011 | 24 | healthy | wash | no | adults | wash - saline / swab -VTM, STGG | Culture and PCR | 40 | rayon tipped |
| Mitamura | 2012 | 330 | respiratory Infection | aspirate | no | children and adults | wash - VTM/ swab - STGG | DIF | undefined | unknown |

- VTM
Viral Transport Medium - RT-PCR
Reverse-Transcriptase Polymerase Chain Reaction - DIF
Direct Immunoflourescence - STGG
Skim Milk, Tryptone, Glucose, and Glycerin - R-Mix
“R-Mix” cell culture medium (Diagnostic Hybrids, Athens, OH)

### Laboratory methods

Methods used to detect pathogen carriage varied across studies. Eleven employed species-specific molecular methods: seven using PCR and six using direct immunofluorescence, including one using both. Five different immunoassay kits were used for immunofluorescence. Three cultured on inoculated Skim-milk-tryptone-glucose-glycerin for bacteria and one used the “R-mix” rapid culture method for viral growth. Where recorded, the cycle threshold for PCR was >40 with the exception of one which was >35. Six used saline to transport the samples, six used viral transport medium, one used Guianidinium Thiocyanate Buffer and one was unspecified.

### Virology

PCR analysis of NA and NPS for group A pathogens, i.e those associated with hospitalising illness, included Respiratory Syncytial Virus, Parainfluenza virus, Metapneumovirus and Influenza A+B [figure 4]. Sensitivities as a fraction of consensus standard (positive for either collection method) ranged between 84% and 96% for these pathogens by both collection methods. Similar lack of statistical dissimilarity was found when stratifying for group B viruses Rhinovirus, Adenovirus, Coronavirus^*^, Enterovirus (not normally associated with severe disease), but the range of sensitivities were greater [figure 5]^54,55,56,57,58^. When immunofluorescence was utilised for the same, there was no improved sensitivity for NA and NPS, with the exception of Influenza A (H1N1) where swabs had a greater sensitivity (p<0.001)^59,57,60^. Nasal swabs for influenza was similarly demonstrated to have greater sensitivity for detecting Influenza in one study of 122 participants [figure 6]^61^. Another study of 89 paired samples, in which the exact nature of nasal washes was not described in detail, found the exact same RSV and Parainfluenzavirus Influenza sensitivity, and a near equivocal influenza swab/wash sensitivity of 100/97.1 (p-value =1)^62^ Many studies described findings seeming to advantage one collection method or other as a standalone, but this disappeared when pooled with others testing for the same pathogen by the same techniques. Indeed, when combined, the sensitivities of both collection methods for Parainfluenzavirus was the same (84.7%/84.7%).

**Figure 4.**
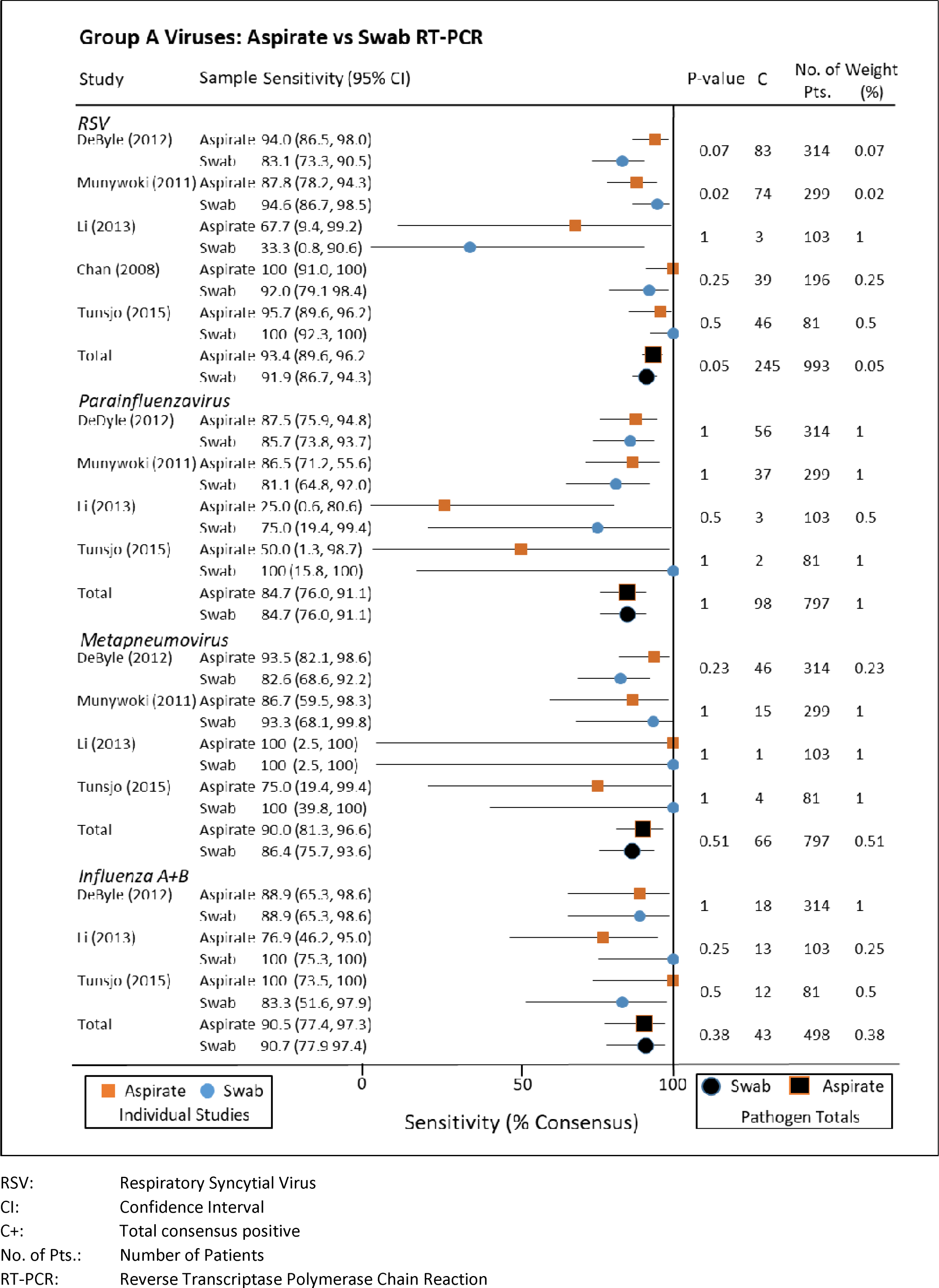
Group A viruses: Aspirates vs. Swab RT-PCR. RSV: Respiratory Syncytial Virus CI: Confidence Interval C+: Total consensus positive No. of Pts.: Number of Patients RT-PCR: Reverse Transcriptase Polymerase Chain Reaction

**Figure 5.**
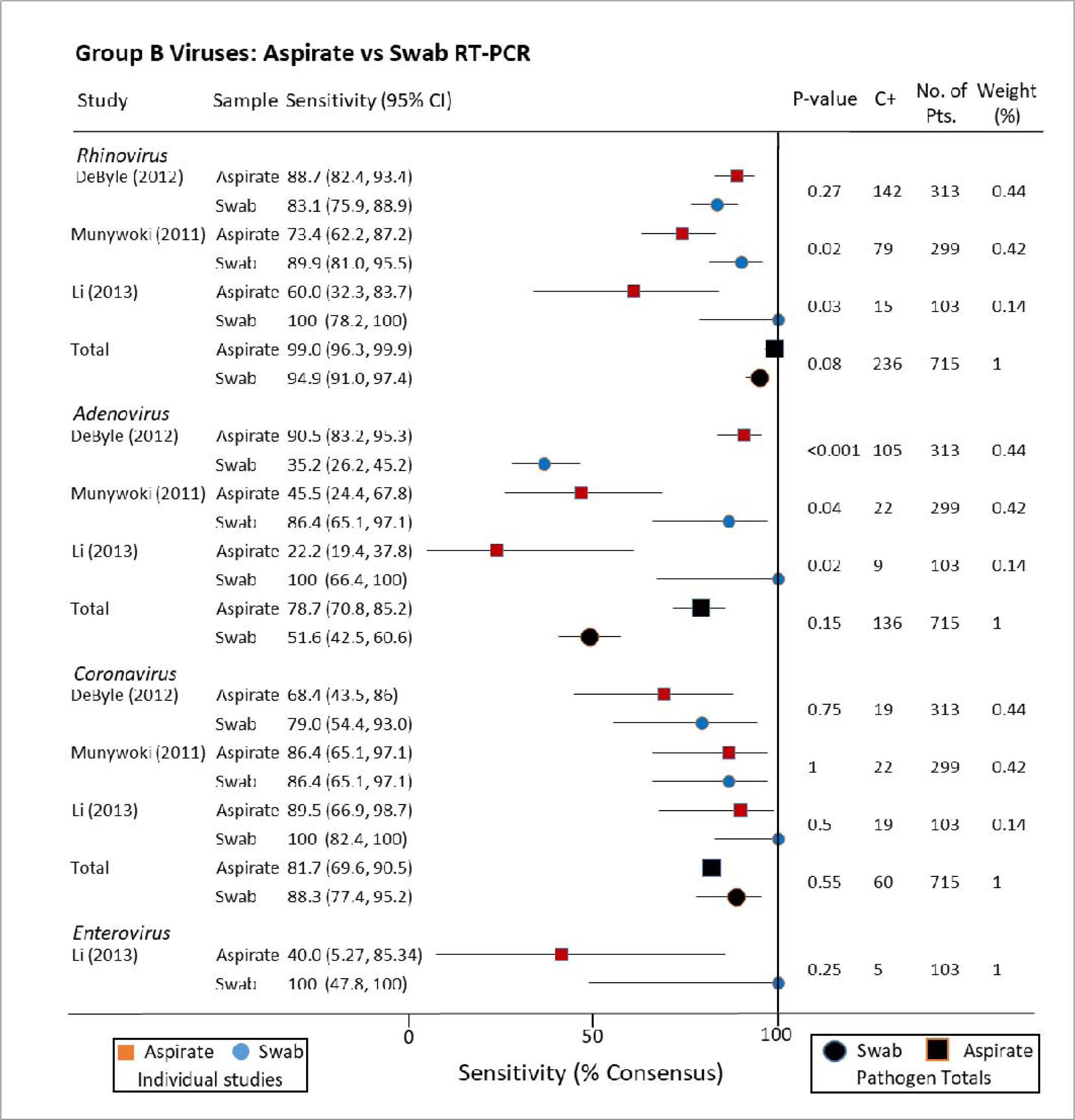
Group B viruses: Aspirates vs. Swab RT-PCR. CI: Confidence Interval C+: Total consensus positive No. of Pts.: Number of Patients RT-PCR: Reverse Transcriptase Polymerase Chain Reaction

**Figure 6.**
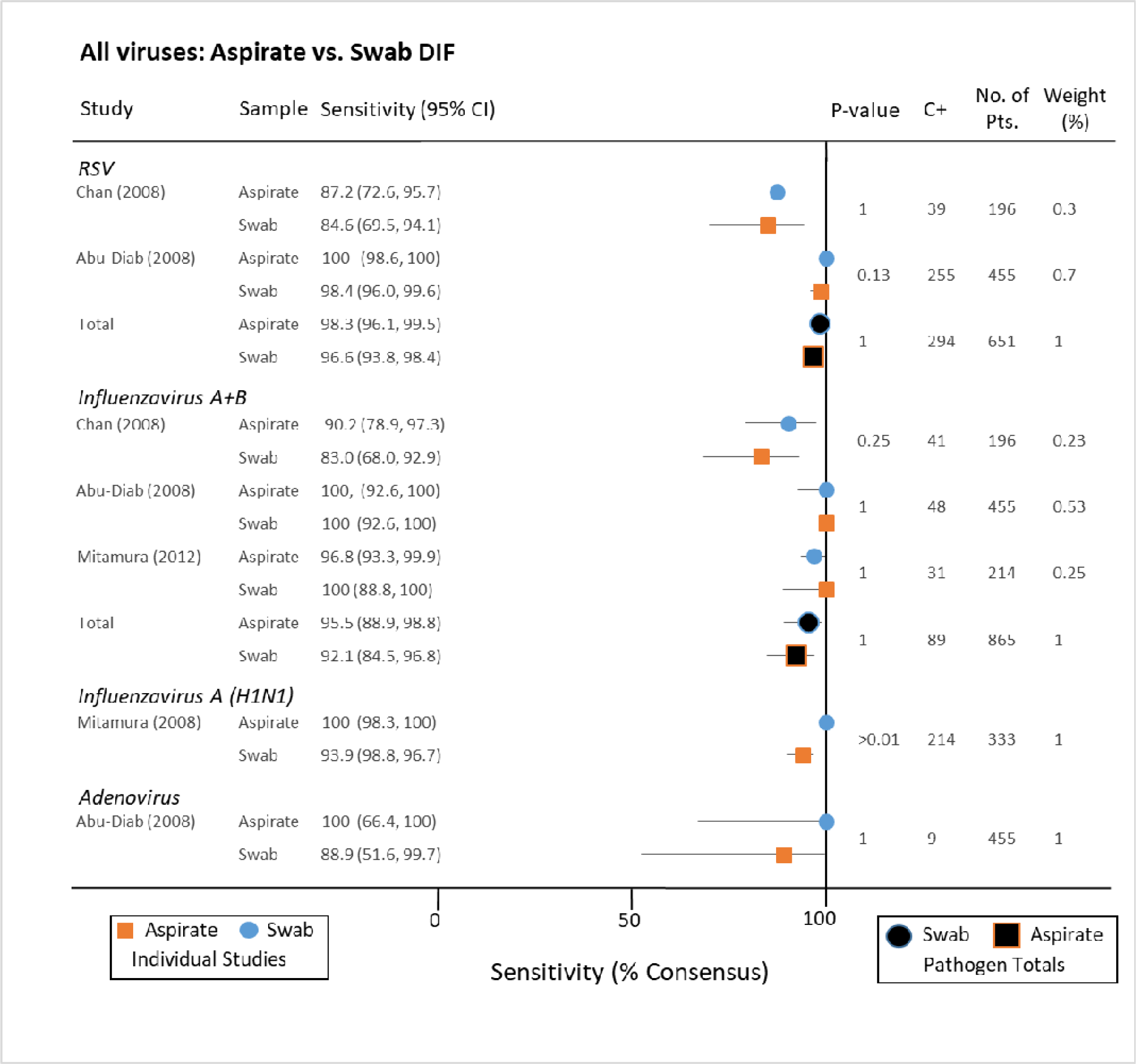
All viruses: Aspirates vs. Swab DIF. RSV: Respiratory Syncytial Virus CI: Confidence Interval C+: Total consensus positive No. of Pts.: Number of Patients DIF: Direct Immunofluorescence

### Bacteriology

Neither aspirate-wash versus swab for *Bordetella pertussis* PCR, nor non-typable *Haemophilus influenzae* in culture, yielded a significant advantage^63,64^. Collated sensitivities of the Naclerio method vs. NFS for a variety of species in 24 healthy British adults favoured NFS for *Neisseria* (60.2/100%), Diptherioids (66.7/100%), and Alpha-haemolytic streptococci (18.8/100%, p <0.001), the Naclerio method for *Staphylococcus aureus* (100/66.7%), and equivocal for *Moraxella catarrhallis* ^*33*^. A similar number of Kenyan infants presenting to hospital with mild illness not requiring hospitalisation, and having a suction catheter passed to the nasopharynx grew strep. pneumoniae in 55 samples. In comparison, 47 (85.0%) of these grew the pathogen on their NFS (p=0.005)^65^. These high yields may reflect the later adoption of the pneumococcal vaccine in Kenya in 2011^66^.

## Discussion

This systematic review found a moderate body of evidence comparing nasopharyngeal swabs with aspirates and washes with no significant difference in sensitivity. These findings were predominantly with PCR, comparing swabs with suction-using aspirates, and covered a range of potential viruses. Beyond these strata data were sparse, particularly for purely wash-based methods, and for detection of bacteria. Statistical significance of higher NP swab sensitivity was high for H1N1, but as this same study found no clear advantage this method for Influenza A nor B, this could be a statistical outlier. Some of the studies included were from an era of DIF and culture, which will have less relevance in future with the predominance of genomic diagnostics. Furthermore, the mechanical understanding at a microscopic level is poorly understood – although swabbing and brushing are more abrasive and likely to access deeper layers of the mucosal barrier. The inherent difference that bacteria and viruses are extra- and intracellular respectively, and the adhesive properties of biofilm also require consideration^67^.

The abundance of confounding variables can only be accounted for in part by the risk of bias assessments. Variations in transport, time in storage and operators persisted, though not deemed to skew the results. Similarly, the strict inclusion criteria for recording of sampling methodology still allowed for much operator dependent intra-and inter-study variability. While a true 100% sensitivity test is elusive, the approximation of similar sensitivity rates for multiple different method comparisons implies saturation of this diagnostic chain: only on the smaller studies were large differences in sensitivities seen, and these not only disappeared when pooling studies, but comparing with better powered studies. This implies a limiting common denominator to all. On the diagnostic chain from mucosa to lab bench, the step least likely to be controlled is the specific anatomical sampling technique. Single operator sampling under direct vision and controlled conditions have been described elsewhere and are required for confidence in the niche sampled^68^. Nevertheless the methodological heterogeneity was controlled by only including case matched control studies, with variables more likely to confound overall than favour one or other sampling method.

Ergonomics also merit consideration: the washing method has been described as more comfortable for adults than a nasopharyngeal swab, and in children anterior nasal swabbing results in a significantly lower infant distress score than an aspirate^33,23^. The perceived and achieved discomfort must also be presumed to affect the thoroughness and accuracy in a linear fashion. In the current pandemic, concern persists around suctioning as an aerosol generating procedure, however swabbing carries its own risk of induced coughing, sneezing or vomiting^69,70^.

### Limitations of current literature

The Cochrane rapid review protocol proved a portable and efficient mode of prompt evidence synthesis for this timely clinical question, purely with the use of open access freeware. This protocol maintains a moderate degree of quality assessment while removing full search saturation and streamlining study selection and data extraction. As rapid reviews are an evolving methodology, it is unclear the extent to which methodological omissions compromise the saturation of these results^71^.

The clinical application of viral detection is not straightforward. Such techniques detect only pathogen carriage and do not demonstrate fulminant respiratory disease. Variations in viral shedding in the upper respiratory tract include: a shorter time to peak viral concentrations in saliva in SARS-CoV-19 compared to severe acute respiratory syndrome (SARS) (5 days vs 7-10 days), and completed viral shedding of Influenzavirus in adults is only completed around 5-7 days compared to infectivity persisting beyond 10 days in infants^72,73,74^. The Attributable Fraction, namely the percentage of times a disease is caused by a detected virus ranges from 12% for Rhinovirus to 93% for Respiratory Syncytial Virus^75^. Thus, even truly reliable results only contribute to a dynamic clinical picture.

Need for better understanding of anatomy of the nose in the literature is also called for. “Nasopharyngeal swabs” are not purely so, as the NP cannot be reached except via contact with the turbinates and septum, thus would be more appropriately named a “pan-nasal” swab. Interdisciplinary collaboration between otolaryngologists, pneumonologists and microbiologists would aid robust education of non-specialists in this regard.

### Future Directions

How else might the sensitivity be improved? Moistening of the swab appears to add little sensitivity to dry swabs^76,77^. Cross sectional audit of health professionals swabbing techniques against WHO guidelines in current pandemic climate could reveal variations in practice and drive quality improvement. A true benchmark for 100% sensitivity will remain elusive, but repeated titres in patients with known disease can give a retrospective estimate of the sensitivity of initial samples.

Future accuracy and resolution in the area of 16S-RNA and whole genome sequencing are vulnerable to conflicting results and applicability concerns from any sampling errors. If there is no clear advantage between collection methods for dichotomous carriage/non-carriage, this still leaves the issue of the best method for describing ecology as a whole. For instance, increased diversity of microbiota are removed from brushing the inferior turbinate compared with nasal washing^78^. Given the aforementioned niche-specific diversity, it is difficult to assess if such variations denote a different topographical area being sampled or a different constellation of organisms preferentially collected by that method, perhaps by being less mucosally adhesive. Detailed delineation of the correctly sampled microbiome is a prerequisite to the future application of such technologies to diagnostics and therapy. Measurements of overall nasal diversity or key operational taxonomic units under controlled conditions may aid establishment of symptom scores for upper respiratory infections which would bypass the need for accurate sampling in the clinic^79,80^. These in turn could aid safe de-escalation of treatment^81,82^. Emerging point of care diagnostics are in readiness to accelerate all accurate and reliable respiratory pathogen sampling to guide timely treatment, and surveillance on a global level^83,84^.

## Data Availability

Additional data such as search terms and protocol can be accessed on PROSPERO https://www.crd.york.ac.uk/prospero/ reference number CRD42020189577

## List of Abbreviations and Acronyms

PRISMA: Preferred Reporting Items for Systematic Reviews and Meta-Analyses
QUADAS-2: Quality Assessment of Diagnostic Accuracy Studies -2
(RT-)(q-)PCR: (Reverse Transcriptase)(quantitative)Polymerase Chain Reaction
SARS(-CoV-19): Severe Acute Respiratory Syndrome(–coronavirus-2019)
DIF: Direct Immunoflourescence
NPS: Nasopharyngeal Swab
NTL: Naso-tragal Length
NA: Nasal Aspiration
NW: Nasal Wash
PROSPERO: International prospective register of systematic reviews
WHO: World Health Organisation
Rayyan QCRI: Rayyan Qatar Computing Research Institute
NP: Nasopharyngeal
RevMan: Review Manager
16S-RNA: 16S - Ribonucleic Acid

## DECLARATIONS

### Competing interests

None to declare

### Funding

None to Declare

### Authors’ contributions

MF Conceived the study, registered the protocol, conducted the literature search, screened titles abstracts and full texts, conducted the risk of bias assessments, performed the statistical analysis and authored the manuscript.

MK Helped provided advice on inclusion and exclusion criteria, provided clinical relevance advice, and reviewed the final manuscript

JD Dual screened a proportion of titles, abstracts and full texts, validated the risk of bias assessments and reviewed the final manuscript

All authors have read and approved the final manuscript

## Acknowledgements

Many thanks to Mr Lucasz Zygan for lending his expertise in Otolaryngology

## Availability of data and materials

Full data, including inclusion and exclusion criteria and search terms can be accessed on the PROSPERO register registration no. CRD42020189577 https://www.crd.york.ac.uk/PROSPERO

## Footnotes

* These studies predate the SARS-COVID-19 strain

## Notes

### Competing Interest Statement

The authors have declared no competing interest.

### Clinical Trial

CRD42020189577

### Funding Statement

No part of this work was supported by funds from any external institutions.

### Author Declarations

Discussed with Ulster University Research Governance Committee and not deemed necessary to undertake ethical review.

## References

1 Information on Rapid Molecular Assays, RT-PCR, and other Molecular Assays for Diagnosis of Influenza Virus Infection [Internet]. Centers for Disease Control and Prevention, National Center for Immunization and Respiratory Diseases (NCIRD). 2019 [cited 1 October 2020]. Available from: https://www.cdc.gov/flu/professionals/diagnosis/molecular-assays.htm

2 Fields B, House B, Klena J, Waboci L, Whistler T, Farnon E. Role of Global Disease Detection Laboratories in Investigations of Acute Respiratory Illness. Journal of Infectious Diseases. 2013;208(suppl 3):S173–S176.

3 Surveillance strategies for COVID-19 human infection: Interim Guidance [Internet]. World Health Organisation; 2020 [cited 1 October 2020]. Available from: https://www.euro.who.int/en/health-topics/health-emergencies/coronavirus-covid-19/technical-guidance/2020/surveillance-strategies-for-covid-19-human-infection-interim-guidance,-10-may-2020

4 WHO recommendations on the use of rapid testing for influenza diagnosis [Internet]. World Health Organisation; 2004 [cited 1 October 2020]. Available from: https://www.who.int/influenza/resources/documents/rapid_testing/en/

5 Fang Y, Zhang H, Xie J, Lin M, Ying L, Pang P, Ji W. Sensitivity of Chest CT for COVID-19: Comparison to RT-PCR. Radiology. 2020 Aug;296(2):E115–E117.

6 6Chartrand C, Tremblay N, Renaud C, Papenburg J. Diagnostic Accuracy of Rapid Antigen Detection Tests for Respiratory Syncytial Virus Infection: Systematic Review and Meta-analysis. J Clin Microbiol. 2015 Dec;53(12):3738–49.

7 Souf S. Recent advances in diagnostic testing for viral infections. Bioscience Horizons. 2016;9(hzw010).

8 Steininger C, Kundi M, Aberle SW, Aberle JH, Popow-Kraupp T. Effectiveness of reverse transcription-PCR, virus isolation, and enzyme-linked immunosorbent assay for diagnosis of influenza A virus infection in different age groups. J Clin Microbiol. 2002 Jun;40(6):2051–6.

9 Kuypers J, Wright N, Ferrenberg J, Huang ML, Cent A, Corey L, Morrow R. Comparison of real-time PCR assays with fluorescent-antibody assays for diagnosis of respiratory virus infections in children. J Clin Microbiol. 2006 Jul;44(7):2382–8.

10 Zou L, Ruan F, Huang M, Liang L, Huang H, Hong Z, Yu J, Kang M, Song Y, Xia J, Guo Q, Song T, He J, Yen HL, Peiris M, Wu J. SARS-CoV-2 Viral Load in Upper Respiratory Specimens of Infected Patients.N Engl J Med. 2020 Mar 19;382(12):1177–1179.

11 Mawaddah A, Gendeh HS, Lum SG, Marina MB. Upper respiratory tract sampling in COVID-19. Malays J Pathol. 2020 Apr;42(1):23–35.

12 Callahan C, Lee R, Lee G, Zulauf KE, Kirby JE, Arnaout R. Nasal-Swab Testing Misses Patients with Low SARS-CoV-2 Viral Loads. medRxiv [Preprint]. 2020 Jun 14:2020.06.12.20128736.

13 Luna PN, Hasegawa K, Ajami NJ, Espinola JA, Henke DM, Petrosino JF, Piedra PA, Sullivan AF, Camargo CA Jr, Shaw CA, Mansbach JM. The association between anterior nares and nasopharyngeal microbiota in infants hospitalized for bronchiolitis. Microbiome. 2018 Jan 3;6(1):2.

14 Satzke C, Turner P, Virolainen-Julkunen A, Adrian PV, Antonio M, Hare KM, Henao-Restrepo AM, Leach AJ, Klugman KP, Porter BD, Sá-Leão R, Scott JA, Nohynek H, O’Brien KL; WHO Pneumococcal Carriage Working Group. Standard method for detecting upper respiratory carriage of Streptococcus pneumoniae: updated recommendations from the World Health Organization Pneumococcal Carriage Working Group. Vaccine. 2013 Dec 17;32(1):165–79.

15 Unilateral Nasal Floor and Inferior Meatus Flap [Internet]. Ento Key. 2018 [cited 1 October 2020]. Available from: https://entokey.com/unilateral-nasal-floor-and-inferior-meatus-flap/

16 Gizurarson S. Anatomical and histological factors affecting intranasal drug and vaccine delivery. Curr Drug Deliv. 2012 Nov;9(6):566–82.

17 Liu T, Li N, Dong N. How to Obtain a Nasopharyngeal Swab Specimen. N Engl J Med. 2020 Jul 16;383(3):e14. doi: 10.1056/NEJMc2015949.

18 Wang TC, Kuo LL, Lee CY. Utilizing nasal-tragus length to estimate optimal endotracheal tube depth for neonates in Taiwan. Indian J Pediatr. 2011 Mar;78(3):296–300.

19 Gray MM, Delaney H, Umoren R, Strandjord TP, Sawyer T. Accuracy of the nasal-tragus length measurement for correct endotracheal tube placement in a cohort of neonatal resuscitation simulators. J Perinatol. 2017 Aug;37(8):975–978.

20 Carver C, Jones N. Comparative accuracy of oropharyngeal and nasopharyngeal swabs for diagnosis of COVID-19 [Internet]. Oxford: Centre for Evidence Based Medicine; 2020 [cited 1 October 2020]. Available from: https://www.cebm.net/covid-19/comparative-accuracy-of-oropharyngeal-and-nasopharyngeal-swabs-for-diagnosis-of-covid-19/

21 Spencer S, Thompson MG, Flannery B, Fry A. Comparison of Respiratory Specimen Collection Methods for Detection of Influenza Virus Infection by Reverse Transcription-PCR: a Literature Review. J Clin Microbiol. 2019 Aug 26;57(9):e00027–19.

22 LeBlanc JJ, Heinstein C, MacDonald J, Pettipas J, Hatchette TF, Patriquin G. A combined oropharyngeal/nares swab is a suitable alternative to nasopharyngeal swabs for the detection of SARS-CoV-2. J Clin Virol. 2020 Jul;128:104442.

23 Macfarlane P, Denham J, Assous J, Hughes C. RSV testing in bronchiolitis: which nasal sampling method is best? Arch Dis Child. 2005 Jun;90(6):634–5.

24 Chan PK, To WK, Ng KC, Lam RK, Ng TK, Chan RC, Wu A, Yu WC, Lee N, Hui DS, Lai ST, Hon EK, Li CK, Sung JJ, Tam JS. Laboratory diagnosis of SARS. Emerg Infect Dis. 2004 May;10(5):825–31.

25 Public Health England. How to use the self-swabbing kit for a combined throat and nose swab (video) [Internet]. 2020 [cited 1 October 2020]. Available from: https://www.youtube.com/watch?v=5qHTBlxfNes&feature=youtu.be

26 Ek P, Böttiger B, Dahlman D, Hansen KB, Nyman M, Nilsson AC. A combination of naso-and oropharyngeal swabs improves the diagnostic yield of respiratory viruses in adult emergency department patients. Infect Dis (Lond). 2019 Apr;51(4):241–248.

27 Kim C, Ahmed JA, Eidex RB, Nyoka R, Waiboci LW, Erdman D, Tepo A, Mahamud AS, Kabura W, Nguhi M, Muthoka P, Burton W, Breiman RF, Njenga MK, Katz MA. Comparison of nasopharyngeal and oropharyngeal swabs for the diagnosis of eight respiratory viruses by real-time reverse transcription-PCR assays. PLoS One. 2011;6(6):e21610.

28 Daley P, Castriciano S, Chernesky M, Smieja M. Comparison of flocked and rayon swabs for collection of respiratory epithelial cells from uninfected volunteers and symptomatic patients. J Clin Microbiol. 2006 Jun;44(6):2265–7.

29 Bruijns BB, Tiggelaar RM, Gardeniers H. The Extraction and Recovery Efficiency of Pure DNA for Different Types of Swabs. J Forensic Sci. 2018 Sep;63(5):1492–1499.

30 Esposito S, Molteni CG, Daleno C, Valzano A, Cesati L, Gualtieri L, Tagliabue C, Bosis S, Principi N. Comparison of nasopharyngeal nylon flocked swabs with universal transport medium and rayon-bud swabs with a sponge reservoir of viral transport medium in the diagnosis of paediatric influenza. J Med Microbiol. 2010 Jan;59(Pt 1):96–99.

31 Moore C, Corden S, Sinha J, Jones R. Dry cotton or flocked respiratory swabs as a simple collection technique for the molecular detection of respiratory viruses using real-time NASBA. J Virol Methods. 2008 Nov;153(2):84–9.

32 WHO guidelines for the collection of human specimens for laboratory diagnosis of avian influenza infection [Internet]. World Health Organisation; 2005 [cited 1 October 2020]. Available from: https://www.who.int/influenza/human_animal_interface/virology_laboratories_and_vaccines/guidelines_collection_h5n1_humans/en/

33 Gritzfeld JF, Roberts P, Roche L, El Batrawy S, Gordon SB. Comparison between nasopharyngeal swab and nasal wash, using culture and PCR, in the detection of potential respiratory pathogens. BMC Res Notes. 2011 Apr 13;4:122.

34 Leach AJ, Stubbs E, Hare K, Beissbarth J, Morris PS. Comparison of nasal swabs with nose blowing for community-based pneumococcal surveillance of healthy children. J Clin Microbiol. 2008 Jun;46(6):2081–2.

35 van den Bergh MR, Bogaert D, Dun L, Vons J, Chu ML, Trzciński K, Veenhoven RH, Sanders EA, Schilder AM. Alternative sampling methods for detecting bacterial pathogens in children with upper respiratory tract infections. J Clin Microbiol. 2012 Dec;50(12):4134–7.

36 Tang S, Hemyari P, Canchola JA, Duncan J. Dual composite reference standards (dCRS) in molecular diagnostic research: A new approach to reduce bias in the presence of Imperfect reference. J Biopharm Stat. 2018;28(5):951–965.

37 Umemneku Chikere CM, Wilson K, Graziadio S, Vale L, Allen AJ. Diagnostic test evaluation methodology: A systematic review of methods employed to evaluate diagnostic tests in the absence of gold standard - An update. PLoS One. 2019 Oct 11;14(10):e0223832.

38 Worster A, Carpenter C. Incorporation bias in studies of diagnostic tests: how to avoid being biased about bias. CJEM. 2008 Mar;10(2):174–5.

39 MERS-CoV | Interim Guidelines for Clinical Specimens from PUI | CDC [Internet]. 2020 [cited 1 October 2020]. Available from: https://www.cdc.gov/coronavirus/mers/guidelines-clinical-specimens.html

40 Ek P, Böttiger B, Dahlman D, Hansen KB, Nyman M, Nilsson AC. A combination of naso- and oropharyngeal swabs improves the diagnostic yield of respiratory viruses in adult emergency department patients. Infect Dis (Lond). 2019 Apr;51(4):241–248.

41 Arevalo-Rodriguez I, Buitrago-Garcia D, Simancas-Racines D, Zambrano-Achig P, del Campo R, Ciapponi A, Sued O, Martinez-Garcia L, Rutjes A, Low N, Bossuyt PM, Perez-Molina JA, Zamora A. False-negative results of initial RT-PCR assays for COVID-19: A Systematic Review. medRxiv. 2020;20066787(doi:10.1101/2020.04.16.20066787%J).

42 Woloshin S, Patel N, Kesselheim AS. False Negative Tests for SARS-CoV-2 Infection -Challenges and Implications. N Engl J Med. 2020 Aug 6;383(6):e38.

43 Tricco A, Langlois E, Strauss S. Rapid reviews to strengthen health policy and systems: a practical guide. World Health Organization Alliance for Health Policy and Systems Research; 2017.

44 Tricco AC, Antony J, Zarin W, Strifler L, Ghassemi M, Ivory J, Perrier L, Hutton B, Moher D, Straus SE. A scoping review of rapid review methods. BMC Med. 2015 Sep 16;13:224.

45 Garritty C, Gartlehner G, Kamel C, King V, Nussbaumer-Streit B, Stevens A. Cochrane Rapid Reviews. Interim Guidance from the Cochrane Rapid Reviews Methods Group. 2020.

46 Moher D, Shamseer L, Clarke M, Ghersi D, Liberati A, Petticrew M, Shekelle P, Stewart LA; PRISMA-P Group. Preferred reporting items for systematic review and meta-analysis protocols (PRISMA-P) 2015 statement. Syst Rev. 2015 Jan 1;4(1):1.

47 Deeks JJ, Wisniewski S, Davenport C. Chapter 4: Guide to the contents of a Cochrane Diagnostic Test Accuracy Protocol. In: Deeks JJ, Bossuyt PM, Gatsonis C (editors), Cochrane Handbook for Systematic Reviews of Diagnostic Test Accuracy Version 1.0.0. The Cochrane Collaboration, 2013. Available from: http://srdta.cochrane.org/.

48 Booth A, Clarke M, Dooley G, Ghersi D, Moher D, Petticrew M, Stewart L. The nuts and bolts of PROSPERO: an international prospective register of systematic reviews. Syst Rev. 2012 Feb 9;1:2.

49 Ouzzani M, Hammady H, Fedorowicz Z, Elmagarmid A. Rayyan-a web and mobile app for systematic reviews. Syst Rev. 2016 Dec 5;5(1):210.

50 Schoonjans F. MedCalc’s Diagnostic test evaluation calculator [Internet]. MedCalc. 2020 [cited 2 October 2020]. Available from: https://www.medcalc.org/calc/diagnostic_test.php

51 McNemar test on paired proportions [Internet]. SciStat. 2020 [cited 2 October 2020]. Available from: https://www.scistat.com/statisticaltests/mcnemar.php

52 Whiting PF, Rutjes AW, Westwood ME, Mallett S, Deeks JJ, Reitsma JB, Leeflang MM, Sterne JA, Bossuyt PM; QUADAS-2 Group. QUADAS-2: a revised tool for thequality assessment of diagnostic accuracy studies. Ann Intern Med. 2011 Oct 18;155(8):529–36.

53 Review Manager 5 (RevMan 5). The Cochrane Collaboration, 2014. Available from revman.cochrane.org

54 Debyle C, Bulkow L, Miernyk K, Chikoyak L, Hummel KB, Hennessy T, Singleton R. Comparison of nasopharyngeal flocked swabs and nasopharyngeal wash collection methods for respiratory virus detection in hospitalized children using real-time polymerase chain reaction. J Virol Methods. 2012 Oct;185(1):89-93. doi: 10.1016/j.jviromet.2012.06.009. Epub 2012 Jun21. PMID: 22728277; PMCID: PMC4655598.

55 Munywoki PK, Hamid F, Mutunga M, Welch S, Cane P, Nokes DJ. Improved detection of respiratory viruses in pediatric outpatients with acute respiratory illness by real-time PCR using nasopharyngeal flocked swabs. J Clin Microbiol. 2011 Sep;49(9):3365–7. doi: 10.1128/JCM.02231-10. Epub 2011 Jul 20. PMID: 21775539; PMCID: PMC3165583.

56 Li L, Chen QY, Li YY, Wang YF, Yang ZF, Zhong NS. Comparison among nasopharyngeal swab, nasal wash, and oropharyngeal swab for respiratory virus detection in adults with acute pharyngitis. BMC Infect Dis. 2013 Jun 20;13:281. doi: 10.1186/1471-2334-13-281. PMID: 23786598; PMCID: PMC3698019.

57 Chan KH, Peiris JS, Lim W, Nicholls JM, Chiu SS. Comparison of nasopharyngeal flocked swabs and aspirates for rapid diagnosis of respiratory viruses in children. J Clin Virol. 2008 May;42(1):65–9. doi: 10.1016/j.jcv.2007.12.003. Epub 2008 Feb 1. PMID: 18242124.

58 Tunsjø HS, Berg AS, Inchley CS, Røberg IK, Leegaard TM. Comparison of nasopharyngeal aspirate with flocked swab for PCR-detection of respiratory viruses in children. APMIS. 2015 Jun;123(6):473–7. doi: 10.1111/apm.12375. Epub 2015 Apr 23. PMID: 25904242.

59 Mitamura K, Kawakami C, Shimizu H, Abe T, Konomi Y, Yasumi Y, Yamazaki M, Ichikawa M, Sugaya N. Evaluation of a new immunochromatographic assay for rapid identification of influenza A, B, and A(H1N1)2009 viruses. J Infect Chemother. 2013 Aug;19(4):633–8. doi: 10.1007/s10156-012-0533-1. Epub 2012 Dec 20. PMID: 23254398; PMCID: PMC3738839.

60 Abu-Diab A, Azzeh M, Ghneim R, Ghneim R, Zoughbi M, Turkuman S, Rishmawi N, Issa AE, Siriani I, Dauodi R, Kattan R, Hindiyeh MY. Comparison between pernasal flocked swabs and nasopharyngeal aspirates for detection of common respiratory viruses in samples from children. J Clin Microbiol. 2008 Jul;46

61 Agoritsas K, Mack K, Bonsu BK, Goodman D, Salamon D, Marcon MJ. Evaluation of the Quidel QuickVue test for detection of influenza A and B viruses in the pediatric emergency medicine setting by use of three specimen collection methods. J Clin Microbiol. 2006 Jul;44(7):2638–41. doi: 10.1128/JCM.02644-05. PMID: 16825402; PMCID: PMC1489517.

62 Suave L, Doan Q, Steer Q, Book L, Tilley P. Flocked nasopharyngeal swabs vs nasopharyngeal wash for the diagnosis of respiratory viruses in children. Can J Infect Dis Med Microbiol. 2012;23(Suppl 1):53b.

63 Nunes MC, Soofie N, Downs S, Tebeila N, Mudau A, de Gouveia L, Madhi SA. Comparing the Yield of Nasopharyngeal Swabs, Nasal Aspirates, and Induced Sputum for Detection of Bordetella pertussis in Hospitalized Infants. Clin Infect Dis. 2016 Dec 1;63(Suppl 4):S181–S186. doi: 10.1093/cid/ciw521. PMID: 27838671; PMCID: PMC5106614.

64 Winokur PL, Chaloner K, Doern GV, Ferreira J, Apicella MA. Safety and immunological outcomes following human inoculation with nontypeable Haemophilus influenzae. J Infect Dis. 2013 Sep 1;208(5):728–38. doi: 10.1093/infdis/jit238. Epub 2013 May 28. PMID: 23715660; PMCID: PMC3733507.

65 Abdullahi O, Wanjiru E, Musyimi R, Glass N, Scott JA. Validation of nasopharyngeal sampling and culture techniques for detection of Streptococcus pneumoniae in children in Kenya. J Clin Microbiol. 2007 Oct;45(10):3408–10. doi: 10.1128/JCM.01393-07. Epub 2007 Aug 15. PMID: 17699645; PMCID: PMC2045369.

66 Hammitt LL, Etyang AO, Morpeth SC, Ojal J, Mutuku A, Mturi N, Moisi JC, Adetifa IM, Karani A, Akech DO, Otiende M, Bwanaali T, Wafula J, Mataza C, Mumbo E, Tabu C, Knoll MD, Bauni E, Marsh K, Williams TN, Kamau T, Sharif SK, Levine OS, Scott JAG. Effect of ten-valent pneumococcal conjugate vaccine on invasive pneumococcal disease and nasopharyngeal carriage in Kenya: a longitudinal surveillance study. Lancet. 2019 May 25;393(10186):2146–2154.

67 Suh JD, Ramakrishnan V, Palmer JN. Biofilms. Otolaryngol Clin North Am. 2010 Jun;43(3):521-30, viii.

68 Lu YT, Wang SH, Liou ML, Shen TA, Lu YC, Hsin CH, Yang SF, Chen YY, Chang TH. Microbiota Dysbiosis in Fungal Rhinosinusitis. J Clin Med. 2019 Nov 14;8(11):1973.

69 Philpott C, Burrows S. Aerosol-generating procedures in ENT. London: ENT UK; 2020.

70 Infection prevention and control during health care for probable or confirmed cases of Middle East respiratory syndrome coronavirus (MERS-CoV) infection. World Health Organisation; 2019.

71 Gartlehner G, Affengruber L, Titscher V, Noel-Storr A, Dooley G, Ballarini N, König F. Single-reviewer abstract screening missed 13 percent of relevant studies: a crowd-based, randomized controlled trial. J Clin Epidemiol. 2020 May;121:20–28.

72 Wölfel R, Corman VM, Guggemos W, Seilmaier M, Zange S, Müller MA, Niemeyer D, Jones TC, Vollmar P, Rothe C, Hoelscher M, Bleicker T, Brünink S, Schneider J, Ehmann R, Zwirglmaier K, Drosten C, Wendtner C. Virological assessment of hospitalized patients with COVID-2019. Nature. 2020 May;581(7809):465–469.

73 Fiore A, Fry A, Shay D, Gubareva L, Bresee J, Uyeki T. Antiviral Agents for the Treatment and Chemoprophylaxis of Influenza: Recommendations of the Advisory Committee on Immunization Practices (ACIP). Atlanta: National Center for Immunization and Respiratory Diseases; 2020.

74 Lau LL, Cowling BJ, Fang VJ, Chan KH, Lau EH, Lipsitch M, Cheng CK, Houck PM, Uyeki TM, Peiris JS, Leung GM. Viral shedding and clinical illness in naturally acquired influenza virus infections. J Infect Dis. 2010 May 15;201(10):1509–16.

75 Self WH, Williams DJ, Zhu Y, Ampofo K, Pavia AT, Chappell JD, Hymas WC, Stockmann C, Bramley AM, Schneider E, Erdman D, Finelli L, Jain S, Edwards KM, Grijalva CG. Respiratory Viral Detection in Children and Adults: Comparing Asymptomatic Controls and Patients With Community-Acquired Pneumonia. J Infect Dis. 2016 Feb 15;213(4):584–91.

76 Warnke P, Devide A, Weise M, Frickmann H, Schwarz NG, Schäffler H, Ottl P, Podbielski A. Utilizing Moist or Dry Swabs for the Sampling of Nasal MRSA Carriers? An In Vivo and In Vitro Study. PLoS One. 2016 Sep 14;11(9):e0163073.

77 Hagiya H, Mio M, Murase T, Egawa K, Kokumai Y, Uchida T, Morimoto N, Otsuka F, Shiota S. Is wet swab superior to dry swab as an intranasal screening test? J Intensive Care. 2013 Nov 27;1(1):10.

78 Pérez-Losada M, Crandall KA, Freishtat RJ. Two sampling methods yield distinct microbial signatures in the nasopharynges of asthmatic children. Microbiome. 2016 Jun 16;4(1):25.

79 Gopal Krishnan S, Fun WH, Ramadras MD, Yunus R, Lye YF, Sararaks S. Pertussis clinical case definition: Time for change in developing countries? PLoS One. 2019 Jul 10;14(7):e0219534.

80 Taylor JA, Weber WJ, Martin ET, McCarty RL, Englund JA. Development of a symptom score for clinical studies to identify children with a documented viral upper respiratory tract infection. Pediatr Res. 2010;68(3):252–257.

81 Giancola SE, Nguyen AT, L. B, Ahmed O, Higgins C, Sizemore JA, Orwig KW. Clinical utility of a nasal swab methicillin-resistant Staphylococcus aureus polymerase chain reaction test in intensive and intermediate care unit patients with pneumonia. Diagn Microbiol Infect Dis. 2016 Nov;86(3):307–310.

82 Little P, Hobbs FD, Moore M, Mant D, Williamson I, McNulty C, Cheng YE, Leydon G, McManus R, Kelly J, Barnett J, Glasziou P, Mullee M; PRISM investigators. Clinical score and rapid antigen detection test to guide antibiotic use for sore throats: randomised controlled trial of PRISM (primary care streptococcal management). BMJ. 2013 Oct 10;347:f5806.

83 Gunell M, Antikainen P, Porjo N, Irjala K, Vakkila J, Hotakainen K, Kaukoranta SS, Hirvonen JJ, Saha K, Manninen R, Forsblom B, Rantakokko-Jalava K, Peltola V, Koskinen JO, Huovinen P. Comprehensive real-time epidemiological data from respiratory infections in Finland between 2010 and 2014 obtained from an automated and multianalyte mariPOC® respiratory pathogen test. Eur J Clin Microbiol Infect Dis. 2016 Mar;35(3):405–13.

84 Chartrand C, Leeflang MM, Minion J, Brewer T, Pai M. Accuracy of rapid influenza diagnostic tests: a meta-analysis. Ann Intern Med. 2012 Apr 3;156(7):500–11.

